# Individualized Therapy Optimization for Type 2 Diabetes

**DOI:** 10.1101/2025.10.13.25337959

**Authors:** Arina Lozhkina, Camillo Piazza, Zeina Gabr, Maurice Rupp, David Herzig, Lia Bally, André Jaun

## Abstract

**Background and Aims:** Type 2 diabetes is a wide-spread chronic condition in which blood glucose and body weight management constitute essential therapeutic targets. Emerging technologies have the potential to aid complex therapeutic pharmacotherapy choices that are optimally tailored to individual needs. Here we propose an artificial intelligence combining guidelines with clinical features and continuous glucose monitoring (CGM) to optimize therapeutic decision-making.

**Methods:** Therapeutic guidelines are first encoded using a rule-based model and trained on a neural network. Relying on real world evidence outcomes of a specialist outpatient clinic, transfer learning is used to optimize for glucose-lowering therapies that led to successful treatment outcomes defined as an absolute 0.3% reduction in glycated (HbA1c) over 6.5% without increasing body weight for a BMI over 28. Recommendations that deviate from guidelines are described with Shapley values and tested in digital twins for statistical significance. Four CGM-derived glucose-insulin response dynamic factors serve as additional biomarkers.

**Results:** Dual glycemic & weight targets were achieved in actual clinical practice in 51% cases, increasing to 54% when clinical guidelines were followed. Selecting outcomes in the test set that follow individualized recommendations, this increases further to 56% when using only phenotypic markers and to 64% when adding CGM-derived dynamics factors.

**Conclusions:** Tested on the limited patient number available, our findings show that AI can outperform guidelines in complex type 2 diabetes cases by integrating multiple data sources, drawing on experiential clinical insights, and selecting treatments most effective for each patient’s glucose and weight control.

**One-liner:** A neural network is first trained on guidelines and subsequently on real-world evidence outcomes, performing dual glycaemic/weight optimization to improve the management of type 2 diabetes with/out gluco-dynamic parameters extracted from Continuous Glucose Monitoring.

## Introduction

Type 2 diabetes is a multifactorial disease with complex endocrine interactions and comorbidities. When lifestyle modifications are not sufficient, treatment involves glucose-lowering medication using different mechanisms of action, ranging from monotherapy to combination regimens. Given the high prevalence of overweight and obesity in patients with type 2 diabetes [1], glucose-lowering therapies should be selected to optimize both glycaemic control (HbA1c) and body weight in most cases. While clinical guidelines exist [2], multiple regimens are considered appropriate for outcome optimisation, creating challenges in selecting the optimal individualised approach. Recent studies emphasize the need for precision medicine approaches that individualize treatment based on patient phenotypes and therapeutic goals [3].

Several methods have been proposed to guide decision making in T2D. Therapy evaluation is typically designed as emulations of a target trial aimed at population analysis, rather than individual recommendations [4], for example based on decision trees [5], autoregression [6], and gradient boosting methods [7]. In individualized treatment approaches, the clinical context is often limited, and optimization is guided solely by the reduction in hemoglobin A1c [8, 9]. One prototype of decision support in T2D [10] that incorporated dual optimization for both weight and glycaemic control, predicted changes in A1c and weight for selected therapies; however, as the authors noted, clinical trial populations in the study did not fully reflect real-world treatment patterns and the diversity of patient populations.

Continuous glucose monitoring (CGM) provides a granular assessment of glycaemic patterns. By informing individual metabolic states and consequently the effect of treatment strategies, CGM has the potential to contribute to monitoring therapeutic responses in patients with type 2 diabetes [11, 12]. Current research in the field is mostly focused on the stratification of patients to forecast the risk of complications, and generally overlooks the potential to forecast therapeutic effects [13, 14].

This work aims to optimize dual glycaemic and weight control by integrating CGM-derived gluco-dynamics and clinical features into a transfer learning model refined with real-world evidence (RWE).

## Methods

Given the complexity of the decisions to be taken and the relatively limited amount of data to learn, we first develop a rule-based neural network reproducing commonly accepted guidelines with a high degree of accuracy. Historical records with phenotypic data that are available at the time of therapeutic decisions are then used with a transfer learning methodology in conjunction with RWE outcomes to fine-tune the guideline knowledge and to enrich training data with gluco-dynamic markers that were not available to clinicians. Relying on a separate test set, recommendations that deviate from guidelines are explained with Shapley values, defining similar digital twin cohorts so as to ensure that deviations from guidelines are statistically significant in the retrospective analysis.

Clinical guidelines for the management of type 2 diabetes (EASD&ADA [15]) are first enriched with human knowledge from clinical practice and contextualised to align with local reimbursement frameworks. These guidelines are used to formulate a rule-based model in the form of a decision tree relying on 61 input factors (such as age, weight, A1c, BMI, eGFR, intolerance, current therapy) to choose one of 38 therapeutic combinations (such as ‘metformin + GLP-1 RA’) combining 8 active ingredients (such as ‘GLP-1 RA’). Missing features are set to 0, with a sentinel feature added to account for their absence. A feed-forward neural network with a pyramidal layout of (64,43,22) neurons is then trained with synthetic data to reproduce the rules with more than 85% accuracy independently of white noise that is used for 4 factors that will later record CGM-derived gluco-dynamics, but are not described in the guidelines.

The clinical data are extracted from 6715 anonymized outpatient records spanning from 2009 to 2023 that met the following inclusion criteria: a diagnosis of type 2 diabetes, absence of GAD-65 antibodies, no diagnosis of type 1 or type 3 diabetes, recorded weight and A1c hemoglobin measurements at both the start and end of a treatment regimen. The majority of treatment episodes were concentrated in the later years of the dataset, with the highest proportion observed in 2021 and 2022. The resulting dataset included 853 treatment regimens from 533 patients, with an average age of 60±12 years, a BMI of 33±7 kg/m2 and HbA1c 7.8±1.7%, where the values refer to measurements taken within three months prior to, and no later than, the treatment start date. In that cohort, 61% also suffered from hypercholesterolemia, 46% from chronic kidney disease, and 14% from heart failure. The therapies were the following: 41% first line non-insulin glucose lowering treatment, 37% insulin-treated, 16% mono/dual and 6% triple non-insulin glucose-lowering agents. Drug dosages and pharmacological subclasses were not considered in this analysis.

A subset of the records also included CGM data in the absence of therapy, and was further enriched with four state specific gluco-dynamic coefficients (defined as the rate of glucose uptake by insulin-dependent tissues K_g_, net balance between hepatic glucose output and insulin-independent glucose uptake by brain T_g_, apparent first-order clearance rate for insulin K_i_, and endogenous insulin production rate in the presence of glucose V_i_) obtained from a Gauss-Newton fit of Lotka-Volterra equations describing the evolution of glucose dG/dt = -(K_g_ × G × I) + T_g_ + R (t, w) and insulin dI/dt = -(K_i_ × I) + V_i_ × G + R_i0_ subject to the observed peaks corresponding to food intake R (t, w) = ∑_k_ u_k_ M ((t-t_k_)/w, w) [16].

After splitting the data into 597 (train) and 256 (test) sets and labeling factors that are not available in the data, a neural network is trained (Tensorflow [17]) to predict success of A1c reduction, defined as an absolute reduction of 0.3% or greater (for A1c □ 6.5%) without weight gain in patients for a BMI□ 28.

During the training, the accuracy measured against guidelines drops from 85% to 67%, while the dual optimisation forecasting capability gradually increases from 53% to 62%. A game theory concept known as Shapley values [18] and their kernel SHAP approximation [19] are computed and stored for all the predictions, mapping out a disentangled feature space that is used to define digital twins cohorts.

About one third of the individualized recommendations in the test set differ from guideline rules. Each is evaluated for statistical support to make sure that the forecast draws from similar cases that are likely to share similar outcomes rather than unsupported extrapolations (neural network hallucinations). This is carried out by defining a hypersphere centered on the prediction of interest and using a Manhattan distance to count similar cases falling within a specified radius in Shapley value space. The cumulative number first increases quadratically until an inflection point is reached defining a digital twin cohort with similar characteristics. Simple proportion testing is finally used to compare the outcomes of digital twins under different therapies, rejecting a null hypothesis that individualized alternatives offer no benefit over guidelines with a chosen p-value < 0.05.

## Results

Dual glycemic & weight targets were reached in 131 or 51% of the test set; clinicians strictly adhered to the guidelines in 37 cases, for a slight improvement in outcomes increasing to 54%. Selecting among the 256 real outcomes that coincided with the recommendations of the AI, the model achieved the dual optimization target in 64% of cases when including gluco-dynamics parameters, and 56% when those were left out.

Our model followed guideline-based recommendations in 158 decisions involving patients at an early disease stage or with better glycaemic control (53% at therapy initiation, baseline A1c between 7.2 and 7.7% with a 95% confidence) and often resulted in monotherapy (23% GLP-1 RA, 12% SGLT-2).

When the decisions got more complex and could be enriched with CGM-derived gluco-dynamics parameters, the model identified 98 precision medicine recommendations that differed from guidelines, 49 of which had statistically significant digital twins where the no-benefit hypothesis of the alternative over guidelines could be rejected (p-value < 0.05). Figure 1 illustrates how the therapeutic success rate improves between 20 and 50% when switching from guidelines to the alternative precision medicine recommendation. Further enriched with gluco-dynamics, deviations from guidelines even have a 1.5 times greater likelihood of simultaneously reducing A1c hemoglobin and managing weight.

**Figure 1.**
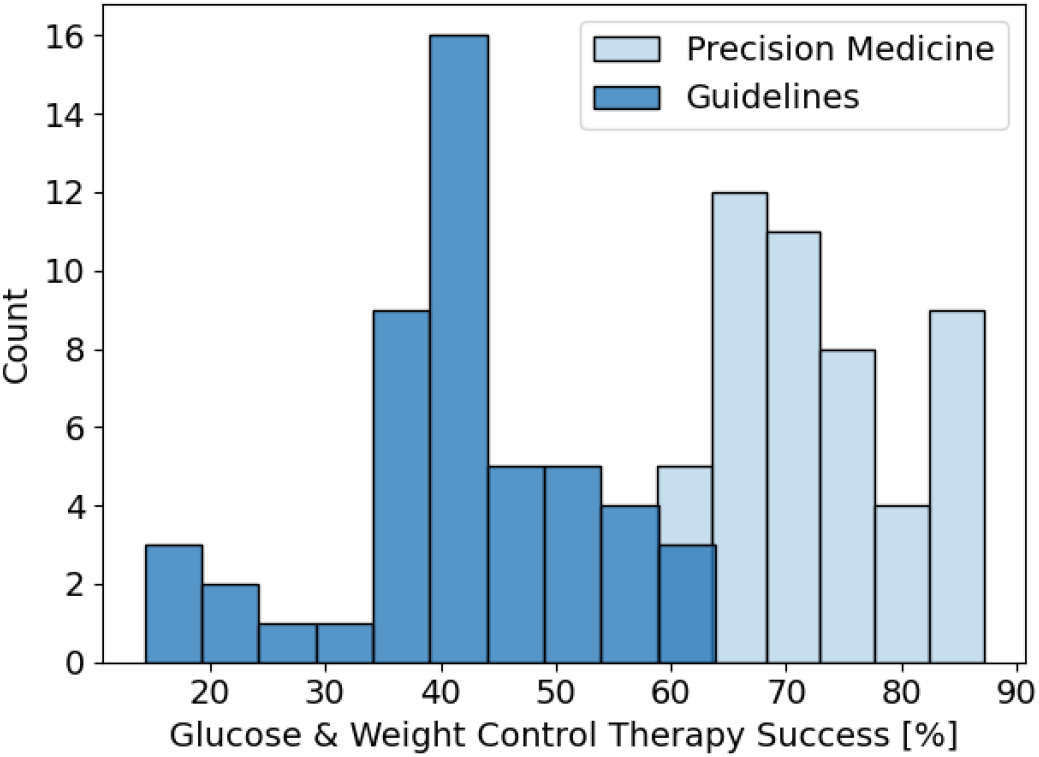
Success rate in achieving dual glycemic & weight control targets when following guideline recommendations versus their statistically significant precision medicine alternative.

The largest group among the 98 therapy reassignments was from GLP-1 RA in combination with insulin to GLP-1 RA alone (19 / 29 statistically significant recommendations). The second largest was switching from insulin to GLP-1 RA (7 / 8 statistically significant). The remaining ones were to replace GLP-1 RA by SGLT-2i in combinations with insulin (6 / 6 statistically significant), and to replace SGLT-2i by metformin in combinations with insulin (6 / 6 statistically significant). To contrast, when gluco-dynamics parameters were artificially switched off, the analysis identified 88 recommendations, 47 of which were statistically significant. Even though our study is limited by sample size, it shows that without gluco-dynamics from CGM data, the tailoring of therapies is reduced, especially for the replacement of GLP-1 RA by SGLT-2i (5 cases) and of SGLT-2i by metformin (3 cases) in combinations with insulin.

The primary advantage of gluco-dynamics parameters (,,, ) over sample statistics such as Glucose Management Indicator (GMI) and Time In Range (TIR, percentage of time glucose values are within the target range of 3.9-10.0 mmol/l) is the timely knowledge of a balance that has long term consequences when decisions are taken close to the boundaries. For example, a patient with 74% TIR and 6.8% A1c hemoglobin will have guidelines recommending metformin, even as the gluco-dynamics parameters indicate high glucose variability that justifies a more intensive treatment with SGLT-2i to stabilize A1c levels and weight.

The most impactful decision that gluco-dynamics parameters bring about concerns a group of patients where the guidelines suggest a combination of metformin, GLP-1 RA plus insulin, while the model suggests to switch the GLP-1 RA with an SGLT-2i with dual optimisation target achievements displayed in Figure 2.

**Figure 2.**
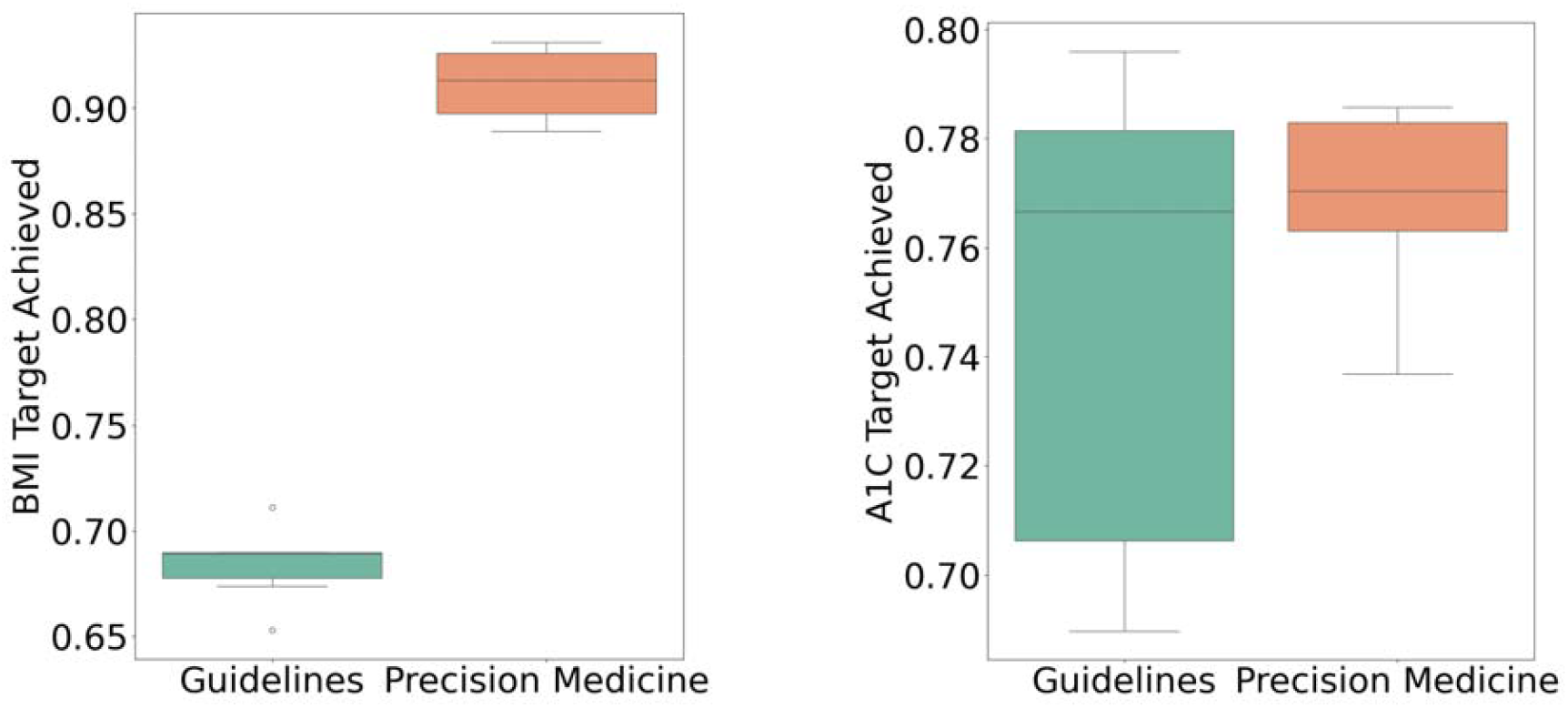
Fraction of 29 guidelines and 19 precision medicine therapies in the digital twin cohort that achieved weight (left) and glycaemic targets (right), in a comparison between those where clinicians followed guideline recommendations GLP-1 RA versus precision medicine SGLT-2i.

A closer look at the Shapley values (not shown in a figure) suggests that when the insulin clearance rate *K*_*i*_ is not high, and that there is no SGLT intolerance, the digital twin signature of individualized reassignments tends to better match patients for whom the guidelines recommend metformin, SGLT-2i and insulin, this however without any simple triplet combination of 23 factors standing out so that it is not possible to distill a simple model.

A more detailed knowledge of gluco-dynamics can noticeably help to deescalate or fine-tune therapies. For example in Figure 3, a patient where the guidelines recommend adding GLP-1 RA with long-acting insulin at 33250 [days] because of a previous rapid-acting insulin therapy, a BMI higher than 28 and a GMI exceeding 7%.

**Figure 3.**
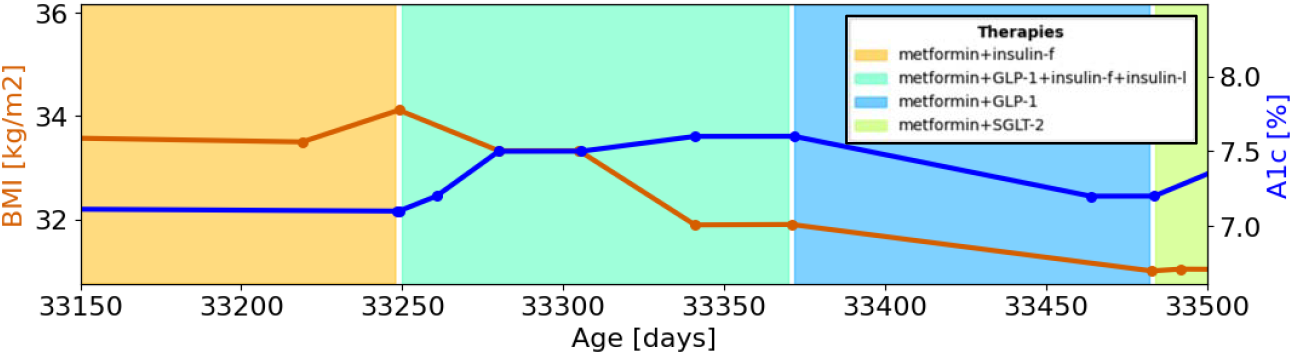
Medical history of a patient^2^ showing the evolution of BMI (in orange, left) and A1c hemoglobin (in blue, right) in the presence of different therapies (colored background) including metformin+insulin (yellow), metformin+GLP1+insulin (turquoise), metformin+GLP1 (blue) and metformin+SGLT-2i (green).

This combination led to a rapid A1c increase from 7.2 to 7.6%, leading a skilled clinician to drop insulin entirely around 33370 [days] effectively stabilizing weight and reducing A1c back to 7.2%. Using only data available at that time, our neural network drew the same conclusions at 33260 [days], justifying the recommendation with sufficient natural insulin production and low insulin clearance, as illustrated by the Shapley values in Figure 4.

**Figure 4.**
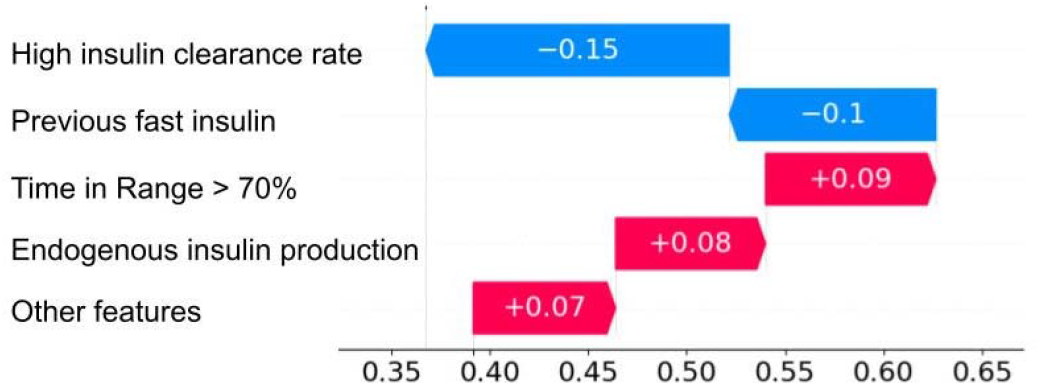
Shapley values explaining how gluco-dynamics such as low *insulin clearance* and sufficient *endogenous insulin production* that can be inferred from CGM leads the neural network to drop insulin from the guidelines recommendation for a patient^3^, leaving only metformin and a GLP-1 RA.

The physiological mechanism is not clearly understood and, in absence of a causal relationship, it is not possible to exclude the possibility that the evolution observed in Figure 3 results from confounding factors that have not been controlled for (e.g. life-style changes, comorbidities, which are absent in Figure 4), neither for the specific patient nor for the statistically significant digital twin cohort that is used here for comparison.

To demonstrate the separation power of gluco-dynamics, consider a therapeutic decision for two patients diagnosed with type 2 diabetes, suffering from obesity (BMI higher than 31), dyslipidemia and chronic kidney disease), having otherwise similar controlled A1c hemoglobin (5.9 and 6%) and medical histories.

Without gluco-dynamics input, the precision medicine model aligns with guidelines for both patients and recommends SGLT-2i. Relying on CGM data available for the first patient and displayed in Figure 5, it is possible to detect a normal insulin sensitivity with an otherwise high endogenous insulin production rate, leading the neural network to switch to an insulin therapy. Again, thanks to a skilled clinician, the switch did happen in reality with a dramatic A1c hemoglobin decrease from 14% to 7.9% and BMI dropping from 31 to 28. In spite of current guidelines, it turns out that the second patient was also put on insulin with a stable BMI and a slight increase of A1c hemoglobin from 6.1 to 6.8% over one year.

**Figure 5.**
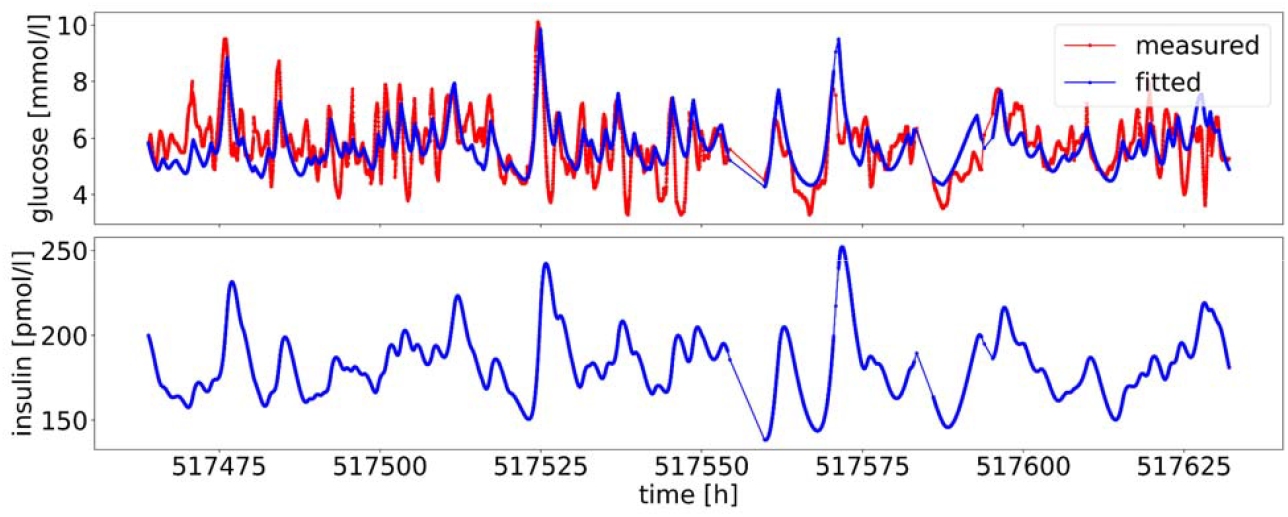
Glucose fitted with an RMSE of 0.84 leading to gluco-dynamic parameters K_g_=0.00174, T_g_=1.30, K_i_=0.747, V_i_=24.9 and insulin generated for the first patient^3^ where the recommendation of the neural network is to switch from SGLT-2i to insulin at time 517632 [hours].

### Clinical evaluation

A usability study has been conducted with 12 general practitioners (GPs) and 12 endocrinology specialists to evaluate how deviations from guidelines are received for 3 groups of deidentified patient records with different clinical profiles and where the AI-generated therapy deviates from guidelines that were current at the time of the evaluation. To maintain a focus on the broader perspective, insulin dosage was pooled into three categories: basal, bolus, or basal-bolus. For each case, the physicians were asked to compare guidelines treatment recommendation with the recommendation from the AI. The differences in opinion, acceptance of AI suggestions, and qualitative feedback were recorded.

Group 1 involved six patients with a mean age of 59.1 years (±9.5), a median BMI of 28.7 kg/m^2^ (interquartile range: 28.7 to 29.6), a mean HbA1c of 7.3% (±0.4), and a median total daily insulin dose (TDD) of 34 units (IQR: 16 to 36). GPs expressed skepticism about replacing GLP1 RA with SGLT-2i (both in combination with metformin and insulin), citing high costs and potential side effects. While some acknowledged the AI’s recommendation as medically sound, the majority preferred continuing GLP-1 RA therapy and stopping insulin cautiously. Specialists generally aligned with these concerns.

Group 2 included seven patients with a mean age of 53.7 years (±10.6), a median BMI of 31.4 kg/m^2^ (IQR: 28.8 to 36.8), a mean HbA1c of 7.6% (±1.1), and a median TDD of 20 units (IQR: 20 to 36). Here, GPs described the AI’s recommendation to replace insulin with SGLT-2i (both in combination with metformin and GLP1 RA) as a useful inspiration but preferred a gradual insulin tapering approach. Some questioned whether the insulin dose was too high to justify a discontinuation of insulin. Specialists agreed and suggested that bariatric surgery might be considered for some patients.

Group 3 involved five patients with a mean age of 66.8 years (±7.2), a median BMI of 28.8 kg/m^2^ (IQR: 28.7 to 35.6), a mean HbA1c of 8.3% (±1.0), and a median TDD of 20 units (IQR: 20 to 55) for whom the guidelines recommended to drop the insulin in a therapy in combination with GLP-1 RA and insulin. In this scenario, GPs were more hesitant to follow the AI’s suggestion to withdraw medications, considering insulin discontinuation appropriate only in palliative contexts. Specialists echoed these reservations, noting that the patients’ HbA1c levels were too elevated to consider stopping insulin altogether.

Two statistical analyses were performed to examine factors influencing acceptance of AI recommendations. First, no statistically significant relationship was found between the total daily insulin dose and the likelihood of accepting the AI-generated suggestion. Second, there was no significant association between the physician’s role (GP vs specialist) and their willingness to follow the AI’s advice. These findings suggest that the decision to accept or reject AI-generated recommendations was not driven by objective metrics such as insulin dosage or by physician specialty.

In summary, the analysis demonstrated that physicians engaged thoughtfully with the AI tool and often viewed its suggestions as clinically reasonable or inspiring. However, final treatment decisions were guided primarily by individual clinical judgment, patient-specific context, and practical considerations such as drug cost, safety, and feasibility. Although the AI system was not routinely followed, its potential to support human decision-making in diabetes management was recognized. The absence of statistically significant predictors for AI acceptance further underscores the nuanced and individualized nature of medical decision-making in clinical practice.

## Conclusion

The complexity of therapeutic decision making in T2D can be overwhelming, even for specialists, and a minority of the decisions are taken in full adherence with clinical guidelines, while only 51% of the therapies achieve dual glycaemic & weight optimisation targets. Learning from real world evidence outcomes in the form of digital twins, this fraction can be improved by individualizing therapies, to 56% with commonly available features and even to 64% when adding gluco-dynamics parameters from CGM recordings.

Trained on guidelines, the algorithm is portable and uses data in a parsimonious fashion to create digital twin cohorts with alternative therapies for which a no benefit hypothesis can be rejected with a suitably low p-value. Personalized treatment proposals consider individual factors including CGM-derived gluco-dynamics parameters, as well as risks and comorbidities, with nuances that may be difficult for a human to incorporate.

A limitation of our work is the lack of personalization of drug dosing and distinctions between subclasses of medications (e.g. different types or dosages of GLP-1RAs) which could not be addressed with the limited amount of data available in this study. In addition, important clinical considerations such as cardiorenal comorbidities (e.g. heart failure) and hypoglycaemia risk were not addressed in this model, despite their established relevance to metabolic treatment in type 2 diabetes. To address these limitations, future work could follow an approach that includes using a larger and more detailed dataset, enhancing the model to incorporate additional medication-related factors, testing the model on diverse patient groups to confirm its accuracy, and finally assessing its real-world effectiveness through a clinical trial. In order to tackle frequently associated comorbidities and complications future research could also incorporate cardiovascular and renal outcomes into the optimization.

In conclusion, we propose a model that integrates clinical guidelines with routine clinical data and CGM-derived gluco-dynamics to identify optimal glucose-lowering therapies for HbA1c and body weight goals for people with type 2 diabetes, demonstrating a promising path toward precision pharmacotherapy.

## Data Availability

This research is based on clinical records that are not openly accessible and cannot be used to identify individuals.

## Acknowledgements

This retrospective study reports research based on clinical records that are not openly accessible and cannot be used to identify individuals. The Ethics committee of Metadvice gave ethical approval for this work. All relevant ethical guidelines have been followed. The work has been supported in part by the Swiss Innovation Agency as Innosuisse project 101.082 IP-LS.

## Notes

### Competing Interest Statement

A.J. is a shareholder of Metadvice

### Author Declarations

Ethics committee of Metadvice gave ethical approval for this work

## References

1. American Diabetes Association. (2025). Standards of care in diabetes—2025. Diabetes Care, 48(Supplement 1), S1–S200. 10.2337/dc25-SINT

2. American Diabetes Association (ADA). Standards of Care in Diabetes—2024. Diabetes Care. 2024;47(Suppl 1). See Section 8: Obesity and Weight Management for the Prevention and Treatment of Type 2 Diabetes, and Section 9: Pharmacologic Approaches to Glycemic Treatment. 10.2337/dc24-S008

3. Wendy K. Chung, Karel Erion, Jose C. Florez, Andrew T. Hattersley, Marie-France Hivert, Christine G. Lee, Mark I. McCarthy, John J. Nolan, Jill M. Norris, Ewan R. Pearson, Louis Philipson, Allison T. McElvaine, William T. Cefalu, Stephen S. Rich, Paul W. Franks; Precision Medicine in Diabetes: A Consensus Report From the American Diabetes Association (ADA) and the European Association for the Study of Diabetes (EASD). Diabetes Care 1 July 2020; 43 (7): 1617–1635. 10.2337/dci20-0022

4. Deng Y, Polley EC, Wallach JD, Herrin J, Ross JS, McCoy RG. Comparative effectiveness of second line glucose lowering drug treatments using real world data: emulation of a target trial. BMJ Med. 2023 Aug 9;2(1):e000419. 10.1136/bmjmed-2022-000419. PMID: 37577025; PMCID: PMC10414064.

5. Ravaut M, Sadeghi H, Leung KK, Volkovs M, Kornas K, Harish V, Watson T, Lewis GF, Weisman A, Poutanen T, Rosella L. Predicting adverse outcomes due to diabetes complications with machine learning using administrative health data. NPJ Digit Med. 2021 Feb 12;4(1):24. 10.1038/s41746-021-00394-8. PMID: 33580109; PMCID: PMC7881135.

6. Tarumi S, Takeuchi W, Qi R, Ning X, Ruppert L, Ban H, Robertson DH, Schleyer T, Kawamoto K. Predicting pharmacotherapeutic outcomes for type 2 diabetes: An evaluation of three approaches to leveraging electronic health record data from multiple sources. J Biomed Inform. 2022 May;129:104001. 10.1016/j.jbi.2022.104001. Epub 2022 Jan 29. PMID: 35101638.

7. Heerspink HJL. Predicting individual treatment response in diabetes. Lancet Diabetes Endocrinol. 2019 Jun;7(6):415–417. 10.1016/S2213-8587(19)30118-4. Epub 2019 Apr 29. PMID: 31047903.

8. Dennis JM, Young KG, Cardoso P, Güdemann LM, McGovern AP, Farmer A, Holman RR, Sattar N, McKinley TJ, Pearson ER, Jones AG, Shields BM, Hattersley AT; MASTERMIND Consortium. A five-drug class model using routinely available clinical features to optimise prescribing in type 2 diabetes: a prediction model development and validation study. Lancet. 2025 Mar 1;405(10480):701–714. 10.1016/S0140-6736(24)02617-5. Epub 2025 Feb 25. PMID: 40020703.

9. Shields BM, Dennis JM, Angwin CD, Warren F, Henley WE, Farmer AJ, Sattar N, Holman RR, Jones AG, Pearson ER, Hattersley AT; TriMaster Study group. Patient stratification for determining optimal second-line and third-line therapy for type 2 diabetes: the TriMaster study. Nat Med. 2023 Feb;29(2):376–383. 10.1038/s41591-022-02120-7. Epub 2022 Dec 7. PMID: 36477733; PMCID: PMC7614216.

10. Buse JB, Holst I, Knop FK, Kvist K, Thielke D, Pratley R. Prototype of an evidence-based tool to aid individualized treatment for type 2 diabetes. Diabetes Obes Metab. 2021 Jul;23(7):1666–1671. 10.1111/dom.14381. Epub 2021 May 4. PMID: 33764641; PMCID: PMC8251774.

11. Klupa T, Czupryniak L, Dzida G, Fichna P, Jarosz-Chobot P, Gumprecht J, Mysliwiec M, Szadkowska A, Bomba-Opon D, Czajkowski K, Malecki MT, Zozulinska-Ziolkiewicz DA. Expanding the Role of Continuous Glucose Monitoring in Modern Diabetes Care Beyond Type 1 Disease. Diabetes Ther. 2023 Aug;14(8):1241–1266. 10.1007/s13300-023-01431-3. Epub 2023 Jun 16. PMID: 37322319; PMCID: PMC10299981.

12. Yoshii H, Mita T, Katakami N, Okada Y, Osonoi T, Aso K, Kurozumi A, Wakasugi S, Sato F, Ishii R, Gosho M, Shimomura I, Watada H. The Importance of Continuous Glucose Monitoring-derived Metrics Beyond HbA1c for Optimal Individualized Glycemic Control. J Clin Endocrinol Metab. 2022 Sep 28;107(10):e3990–e4003. 10.1210/clinem/dgac459. PMID: 35908248; PMCID: PMC9516123.

13. Shao X, Lu J, Tao R, Wu L, Wang Y, Lu W, Li H, Zhou J, Yu X. Clinically relevant stratification of patients with type 2 diabetes by using continuous glucose monitoring data. Diabetes Obes Metab. 2024 Jun;26(6):2082–2091. 10.1111/dom.15512. Epub 2024 Feb 26. PMID: 38409633.

14. Metwally AA, Perelman D, Park H, Wu Y, Jha A, Sharp S, Celli A, Ayhan E, Abbasi F, Gloyn AL, McLaughlin T, Snyder M. Predicting Type 2 Diabetes Metabolic Phenotypes Using Continuous Glucose Monitoring and a Machine Learning Framework. medRxiv [Preprint]. 2024 Sep 9:2024.07.20.24310737. 10.1101/2024.07.20.24310737. PMID: 39108516; PMCID: PMC11302614.

15. European Association for the Study of Diabetes, Statements and Guidelines, https://www.easd.org/guidelines/statements-and-guidelines.html [Accessed 2025]

16. Arina Lozhkina, Zeina Gabr, Camillo Piazza, Maurice Rupp, David Herzig, Lia Bally, André Jaun. Integration of Continuous Glucose Monitoring Dynamics into an AI-based Model for Therapeutic Decision Making in Type 2 Diabetes, Diabetes Technology & Therapeutics Vol. 27, No. S2, 2025, Published 18 March 2025 under https://www.liebertpub.com/doi/10.1089/dia.2024.78502.abstracts.part4b#sec-68

17. Martín Abadi, Ashish Agarwal, Paul Barham, Eugene Brevdo, Zhifeng Chen, Craig Citro, Greg S. Corrado, Andy Davis, Jeffrey Dean, Matthieu Devin, Sanjay Ghemawat, Ian Goodfellow, Andrew Harp, Geoffrey Irving, Michael Isard, Rafal Jozefowicz, Yangqing Jia, Lukasz Kaiser, Manjunath Kudlur, Josh Levenberg, Dan Mané, Mike Schuster, Rajat Monga, Sherry Moore, Derek Murray, Chris Olah, Jonathon Shlens, Benoit Steiner, Ilya Sutskever, Kunal Talwar, Paul Tucker, Vincent Vanhoucke, Vijay Vasudevan, Fernanda Viégas, Oriol Vinyals, Pete Warden, Martin Wattenberg, Martin Wicke, Yuan Yu, and Xiaoqiang Zheng. TensorFlow: Large-scale machine learning on heterogeneous systems, 2015. Software available from https://www.tensorflow.org [Accessed 2025]

18. Shapley, L. 1951. Notes on the end-person game - II. The value of the n-person game. Santa Monica, CA, USA.: RAND Corporation

19. Lundberg S.M. L. S.I. 2017. A unified approach to interpreting model predictions. 31st Conference on Neural Information Processing Systems (NIPS 2017). Long Beach, CA, USA.

